# Do early indicators of euthyroid sick syndrome predict longer-term post-bariatric weight loss? A hypothesis-generating preliminary study

**DOI:** 10.1101/2022.12.15.22283526

**Authors:** Natasha Fowler, Sarah Adler, Thomas Najarian, Carol N. Rowsemitt, Debra L. Safer

**Affiliations:** Department of Behavioral Neuroscience, Oregon Health & Science University, Portland, OR 97239-3098; Department of Psychiatry and Behavioral Sciences, Stanford University School of Medicine, Stanford, CA, USA 94305; Retired, Najarian Center for Obesity, Los Osos, CA, USA 93402; Comprehensive Weight Management, A Nursing Corp., Templeton, CA, USA 93465

## Abstract

**Background:** Despite bariatric surgery’s success for most patients, up to 30% do not experience optimal weight loss outcomes. Reasons remain incompletely understood. Thyroid hormones, due to their role in energy expenditure, have received study. Yet research to date is inconclusive regarding the impact of standard thyroid markers (e.g., thyroid stimulating hormone, or TSH) on weight regulation after bariatric surgery. This prospective observational study investigates whether the early development of euthyroid sick syndrome (ESS) predicts reduced longer-term post-bariatric weight loss. ESS develops during periods of stress (e.g., critical illness, severe infection, famine) and is thought to serve a protective function by suppressing metabolism and conserving weight. Levels of metabolically active free triiodothyronine (FT_3_) drop, levels of non-catabolic reverse 3,3’,5’-triiodothyronine (rT_3_) rise, and TSH levels remain euthyroid. Reductions in the ratio of FT_3_ to rT_3_ significantly predict outcomes in critically ill patients with ESS yet have been unexamined in intentional weight loss among post-bariatric patients. Hence this hypothesis-generating preliminary study investigated whether early changes in the FT_3_:rT_3_ ratio predict weight changes at 1-2 years post-bariatric surgery

**Methods:** Twenty-three adult patients undergoing Roux-en-Y gastric bypass (n=12) or sleeve gastrectomy (n=11) were recruited from a bariatric surgery clinic. The TSH, FT_4_, FT_3_, rT_3_, and self-reported hypothyroid symptoms were collected 2-weeks pre-surgery and between 2 weeks to 3 months post-surgery. Body mass index was measured pre-surgery and 1-2 years post-surgery.

**Results:** Reductions in the FT_3_:rT_3_ ratio from pre- to early post-surgery significantly predicted reduced weight loss at 1 (p= 0.03) and 2 years (p= 0.02) post-surgery. No other thyroid-related markers nor hypothyroid symptoms were predictive.

**Conclusions:** Although replication with larger samples is needed due to sizeable study attrition, this small exploratory study provides provocative support that early changes in the FT_3_:rT_3_ ratio associated with ESS predict longer-term reductions in post-bariatric surgery weight loss. Clinical implications are discussed.

## Introduction

Despite the success of bariatric surgery, up to 30% of patients experience suboptimal weight loss outcomes [1-3]. Reasons for this remain incompletely understood and are likely multifactorial (e.g., behavioral, psychological, surgical technique, physiological factors) [1-4]. Given well-known associations between obesity and impairments in health and quality of life, continued investigations into the identification of putative predictors are needed [1-4].

Metabolic processes interfering with optimal weight loss have received study. Indeed, thyroid hormones, due to their role in energy expenditure, have been identified as possible factors underlying post-bariatric weight regulation [5]. It is well established that thyroid dysfunction, including hyperthyroidism (signaled by an abnormally *low* thyroid stimulating hormone level [TSH]) and hypothyroidism (signaled by an abnormally *high* TSH level) can lead to changes in body weight and resting metabolic rate [5,6]. However, research is inconclusive regarding the contribution of thyroid hormones to weight regulation after bariatric surgery [7,8].

Most of the available research has focused on TSH. These studies show that obese patients who are hypothyroid or euthyroid experience decreases in TSH levels following either bariatric surgery or diet-induced weight loss [7-18]. However, the majority of studies <u>fail</u> to detect a significant relationship between decreases in TSH and the degree of weight loss [8, 11-18]. Other thyroid markers [e.g., triiodothyronine (T_3_), l-thyroxine (T_4_), and T_3_:T_4_], though receiving less study, similarly lack consistent correlations [19]. This lack of association between changes in thyroid hormones and weight loss has been attributed to differences in study samples (e.g., age, gender), variations in the time course of hormone measurements, and/or the use of heterogeneous diet strategies [8,18].

In attempts to control for such inconsistencies, the POUNDS LOST trial [18] examined thyroid hormones and weight loss in response to a consistent diet prescription. Though the study focused on predictive effects of baseline thyroid hormones on weight loss up to 2 years, it also explored correlations between changes in thyroid hormones and weight. No significant association was found for changes in TSH and weight loss.

In contrast, a growing body of evidence documents a consistent pattern regarding thyroid function and the conservation of weight among patients undergoing strict dieting or fasting [20-24]. These changes resemble euthyroid sick syndrome (ESS) [25,26], also known as non-thyroidal illness syndrome or low T_3_ syndrome. ESS is an established response to stresses such as critical illness [21,27-29], non-thyroidal surgery [30], severe infections (including COVID-19 [ 31], as well as famine, anorexia nervosa, and strict hypocaloric dieting [20,22-25].

ESS is generally hypothesized to serve a protective function by lowering the metabolic rate [32,33], hence suppressing weight loss. For example, while levels of the metabolically active thyroid hormone T_3_ /free T_3_ drop, there is a reciprocal rise in the metabolically <u>inactive</u> reverse T_3_ (rT_3_) [25, 26]. Patients remain technically euthyroid, with TSH levels in the low-normal to normal range, without evidence of the abnormally raised TSH levels characteristic of suppressed thyroid function seen in primary hypothyroidism [25,26]. Intriguingly, decreases in the ratio of T_3_ to rT_3_ [or FT_3_ (free T_3_) to rT_3_] robustly predict survival and other key outcomes in patients with ESS, such as in patients with critical illness like fulminant hepatitis [27], non-alcoholic cirrhosis [28], heart failure [29], or those undergoing non-thyroidal surgery [30]. Indeed, the T_3_/rT_3_ (or FT_3_/rT_3_) ratio has been shown to be a better predictor of outcome compared to either of its individual components (e.g., T_3_ or rT_3_) or other measures of thyroid function (e.g., TSH or FT_4_) [30].

No studies to date have examined whether early development of ESS, as signaled by a reduction in the T_3_:rT_3_ ratio, predicts post-bariatric weight loss. Measurement of rT_3_ is rare outside of the ESS literature, given it is traditionally viewed simply as an inactive product of FT_4_ metabolism. [34]. We are aware of only one study, by Lips and colleagues [8], that included measurements of the rT_3_ or the T_3_: rT_3_ ratio in its assessment of the impact of bariatric surgery on thyroid function.

Lips and colleagues reported no correlation between TSH and the degree of weight loss over the 3-month study duration [8]. Interestingly, findings also revealed significant decreases in the T_3_:rT_3_ ratio from baseline to 3 weeks post-surgery, with values of the ratio remaining suppressed through the final 3-month post-surgery follow-up [8]. Such findings are consistent with the diet and famine literature. However, the authors did <u>not</u> investigate whether changes in the T_3_:rT_3_ ratio were correlated with changes in weight. In addition, because data collection ended at 3-month follow-up, the question of whether the observed early decreases in the T_3_:rT_3_ ratio might have predicted later changes in post-bariatric weight loss could not have been determined.

Given the suggestive evidence that rT_3_ plays a pivotal role in suppressing T_3_ production and metabolism in humans [35,36], we examined whether the early development of ESS after intentional weight loss from bariatric surgery, as signaled by a decreased T_3_:rT_3_ ratio, predicts later changes in weight post-surgery. Specifically, this preliminary investigation asks whether reductions in T_3_:rT_3_ between pre-through early post-surgery (e.g., 2 weeks-3 months post-surgery) predict changes in the BMI at 1- and 2-years post-surgery. Exploratory analyses were also included to examine the effects of early changes in hypothyroid symptoms and thyroid-related hormones (e.g., TSH, FT_3_, FT_4_) on later weight loss 1- and 2-years post-surgery.

## Materials and Methods

### Participants

Participants were 23 bariatric surgery patients who were recruited from the Bariatric Surgery and Metabolic Weight Loss Center at a major university medical center between May 1, 2015 and December 31, 2015, when they attended a pre-surgical informational group meeting scheduled 2 weeks pre surgery. The study was described and those who indicated initial interest gave informed written consent and were given more information. All participants were at least 18 years old and met eligibility criteria for either Roux-en-Y gastric bypass (RYGB) or sleeve gastrectomy (SG) [e.g., grade III (BMI ≥ 40 kg/m^2^) or grade II obesity (BMI ≥ 35 and < 40 kg/m^2^ with at least one obesity-related co-morbidity). Patients were excluded from the study if they were prescribed thyroid hormones (e.g., levothyroxine), met criteria for either hypothyroidism or hyperthyroidism pre-surgery as detected by TSH < 0.4 ulU/mL or TSH > 4.0 ulU/mL or FT4 < 0.6 ng/dL or FT4 > 1.6 ng/dl), or did not speak English. All study measures were approved by the Stanford University Institutional Review Board.

The study design included collection of thyroid hormone levels from the hospital clinical laboratory at 2 weeks pre-surgery and early (2 weeks - 3 months) post-surgery. All study-related lab costs were covered by the study. Participants did not receive any study-related compensation for their time completing questionnaires, etc. Post-surgery weight outcomes were collected from the medical chart.

Of the 23 participants with baseline study labs, three had no early post-surgery labs available despite repeated requests by the research team. Of the 20 with both pre- and early post-surgery lab data, four had no weight outcomes in the medical record at 1-year post-surgery and an additional three had no weight outcomes in the medical record at 2-years post-surgery. Reasons for missing 1- or 2-year weight outcomes in the medical record were not available, and baseline demographic differences were explored for those with or without BMI data available at 1- or 2-years post-surgery (see Results).

To examine the impact of early thyroid function on these later weight outcomes, the final data set included participants with these data points. Hence, the final 1-year data set included 16 participants and the final 2-year data set included 13 participants.

### Measures

#### Thyroid-related hormones

Study-related thyroid hormone levels were collected at the university hospital clinical laboratory 2 weeks pre-surgery and early (2 weeks - 3 months) post-surgery to assess effects of early changes in TSH, FT_3_, FT_4_, and rT_3_.

This study chose to assess free T_3_ since it is the form of the hormone available to cells; total T_3_ can be affected by protein binding levels [37]. The ESS research literature utilizes both versions of the ratio (e.g., T3: rT3, FT3: rT3) for practical purposes the T_3_ and FT_3_ decrease in parallel.

The TSH, FT_3_, and FT_4_ levels were assayed by Hillview Laboratory at Stanford University using an electrochemiluminescence immunoassay and rT_3_ levels were assayed by Mayo Clinic Laboratories using liquid chromatography mass spectrometry (LC/MS-MS; for information on the assay specificity and reliability see Zhang et al. [38]. The numerical value of the FT_3_:rT_3_ ratio was calculated using the following formula: FT_3_ (in pg/dL) ÷ rT_3_ (in ng/dL). Reference ranges for each thyroid variable are as follows: TSH = 0.4-4.0 mIU/mL; FT_4_ = 0.6-1.6 ng/dL; FT_3_ = 2.3-4.2 pg/dL; rT_3_ = 9.2-24.1 ng/dL; FT_3_:rT_3_ ≥ 0.20. All laboratory tests were free to study participants.

#### Hypothyroid Symptoms

Patients self-reported whether they were experiencing hypothyroid symptoms at both pre- and post-surgery. Seventeen potential hypothyroid symptoms were queried, including fatigue, sensitivity to cold, irritability, and plateau in weight loss (for the full list of symptoms see Table 1). The total count of endorsed symptoms at each assessment was recorded.

**Table 1.**
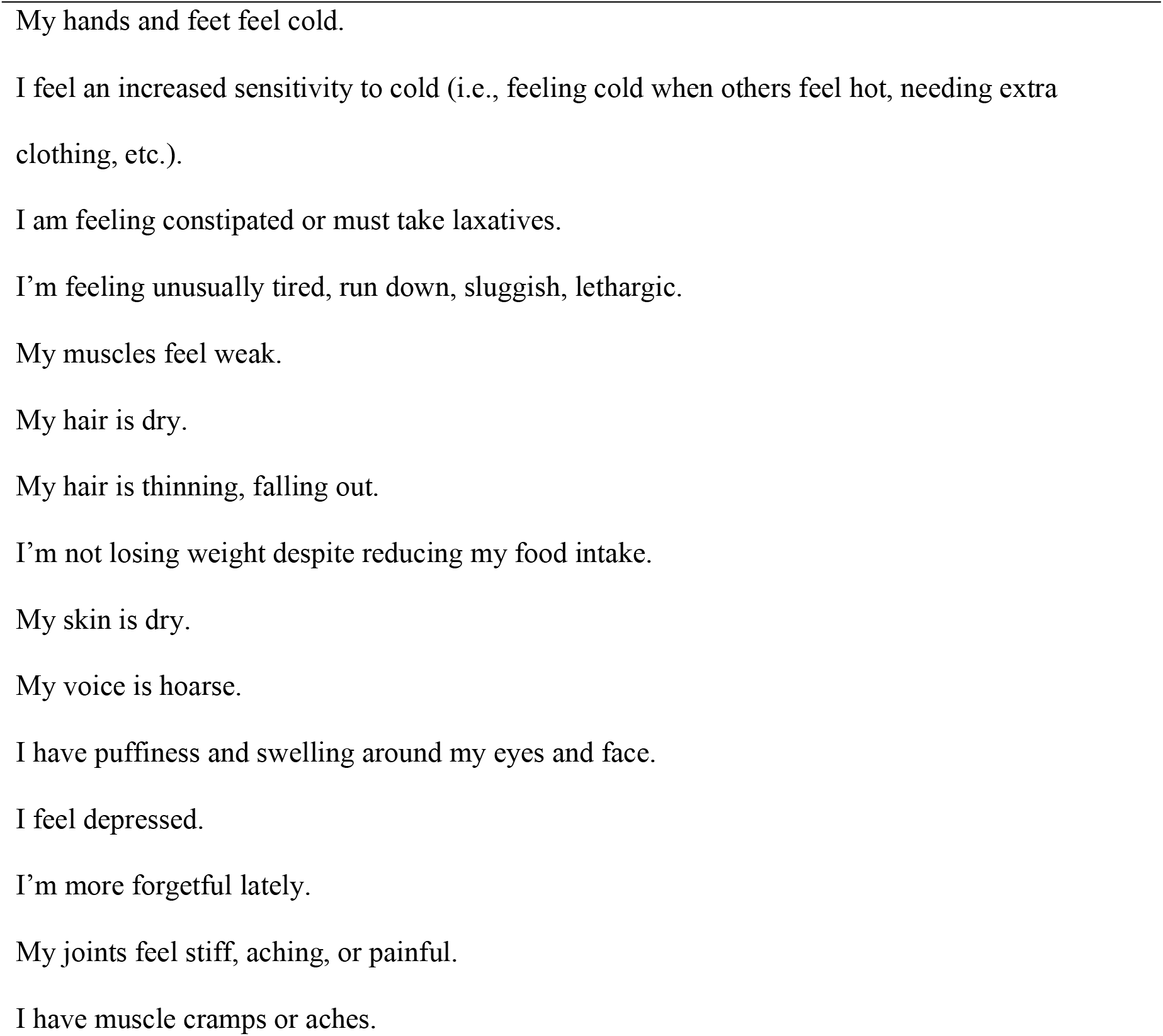
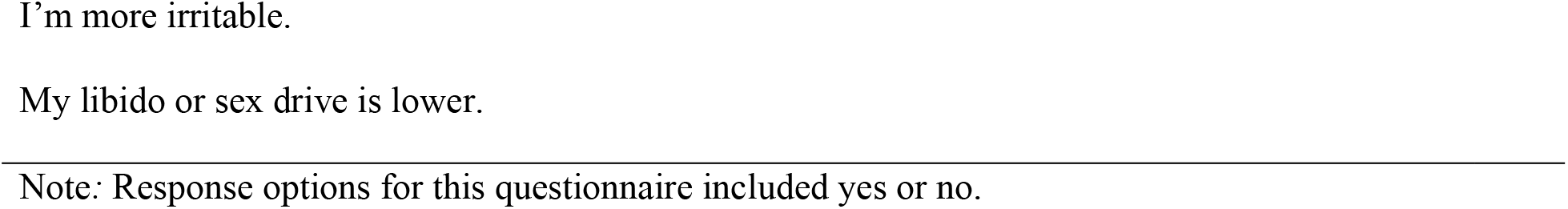
List of Hypothyroid Symptoms.

#### Weight

Participants’ height and weight were measured by the bariatric clinic according to standard procedures. Body weight was assessed on an electronic scale by a trained medical assistant, with the participant in lightweight clothing and shoes removed. Height was measured with a stadiometer in the bariatric clinic. Weight was obtained from the medical record at 1- and 2-years post-surgery. Body Mass Index (BMI) was calculated for each visit as follows: BMI = weight (kilograms)/height^2^ (meters) [39]. Percent of excess BMI loss (% EBMIL) was calculated as follows: % EBMIL = (Initial BMI – Post-surgery BMI)/ (Initial BMI – 25). Ideal BMI was defined, as is standard, as a BMI of 25 kg/m^2^ [39].

### Statistical Analysis

All analyses were conducted using IBM SPSS Statistics for Windows, Version 27.0. 2020. The sum of hypothyroid symptoms experienced pre- and early-post surgery was calculated prior to running analyses. Normality was tested using the Shapiro Wilk test. Non-normally distributed variables were log base 10 transformed and subsequently found to be normal.

Difference scores from pre-surgery to early post-surgery were then calculated for each thyroid variable (e.g., FT_3_:rT_3_ ratio, FT_3_, FT_4_, TSH) and for the total count score for the number of hypothyroid symptoms (see above for rationale for assaying FT_3_ over T_3_). These difference scores were included in analyses as the independent variables. The primary outcome variables were the change in BMI from pre-surgery to 1- and 2-years post-surgery. Individual linear regression models assessed the influence of changes in each thyroid hormone level and hypothyroid symptoms from pre- to early post-surgery on changes in BMI from pre- to 1- and 2-years post-surgery. A surgery moderator variable was included as a covariate in all models to control for effects of surgery type (i.e., RYGB or SG) on weight loss. In addition, Wilcoxon signed rank t-tests were used to assess differences in thyroid hormones and hypothyroid symptom count pre- and early post-surgery. Due to the exploratory nature of this study, corrections were not made for multiple comparisons. A cutoff of p < 0.05 was used during hypothesis testing to indicate significance.

## Results

### Participants

Descriptive statistics are presented in Table 2. Participants in the baseline data set (n = 23) were 91% female, 44.4 (SD = 15.9) years of age, 43.5% Caucasian, 30.4% Hispanic/Latino, and a baseline BMI = 46.2 (SD = 8.5). A little over half the sample (52.2%) underwent Roux-en-Y with the remainder (47.8%) undergoing sleeve gastrectomy. At 1-year post-surgery (n = 16), BMI = 32.6 (SD = 8.1) with % EBMIL = 65.9% (SD = 29.9). At 2-year post-surgery (n = 13), BMI = 31.9 (SD = 6.4) and % EBMIL= 66.4% (SD = 24.9).

**Table 2.**
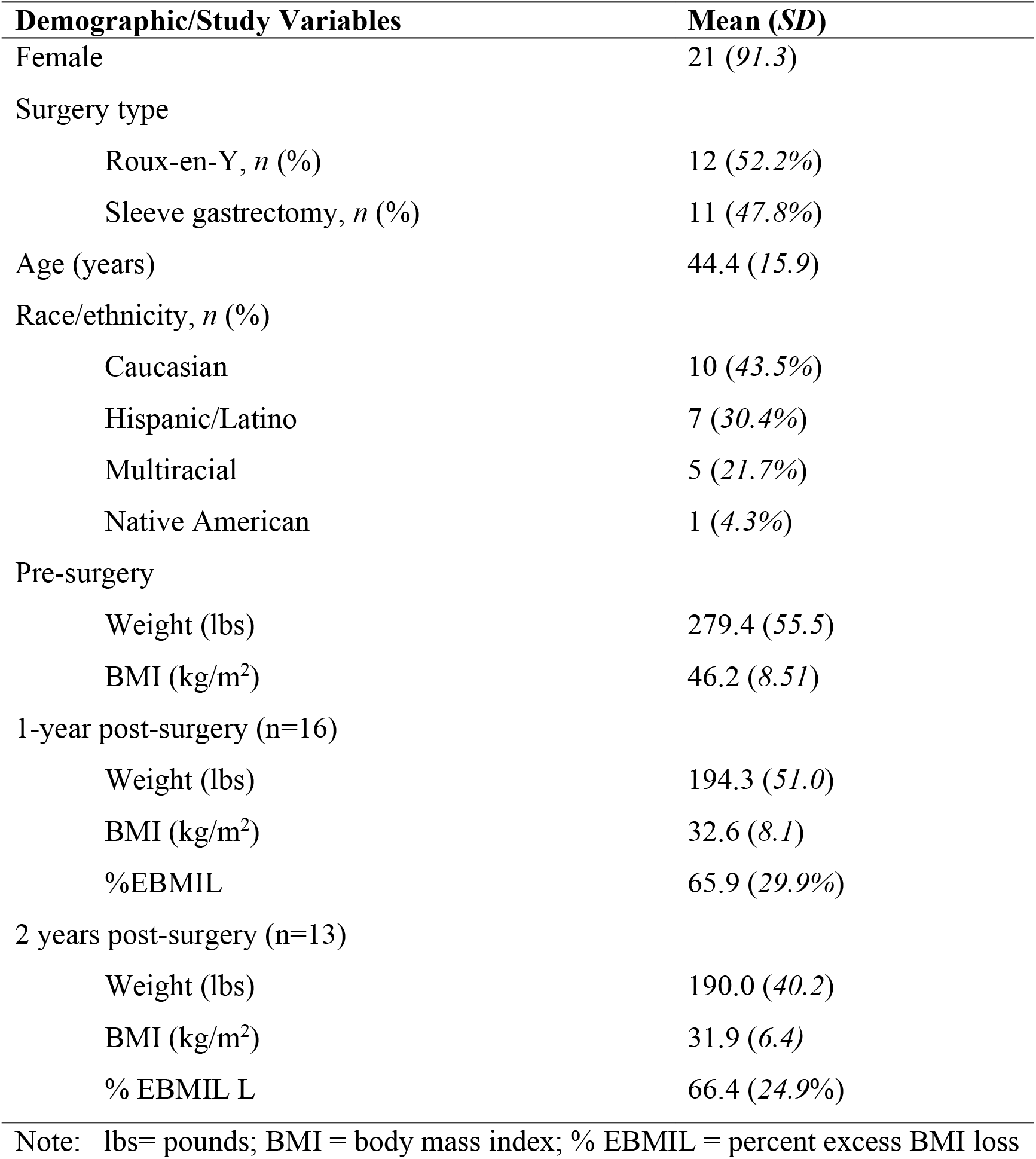
Descriptive data for demographic variables and primary study variables (n = 23)

No significant differences were found on baseline demographic variables (e.g., sex, age, ethnicity, or baseline BMI) between those with or without BMI data available at 1- or 2-years post-surgery.

#### Changes in thyroid parameters and hypothyroid symptoms from baseline to early-post-surgery

Mean values for the individual thyroid hormones and self-reported hypothyroid symptom count. at both the pre-surgery baseline and early (2 weeks - 3 months) post-surgery are presented in Table 3. These findings are germane to the current study only in that the changes from pre- to post-surgery were analyzed as putative predictors of 1-2 year changes in BMI (Table 4).

**Table 3.** Changes in thyroid parameters and hypothyroid symptoms from baseline to early post-surgery (n=20)

| Thyroid Variables | Mean pre-surgery<br>(SD) | Mean early post-surgery<sup>#</sup><br>(SD) | Within normal limits (WNL) at both time points? | *p* |
| --- | --- | --- | --- | --- |
| TSH (mIU/L) | 2.22 (2.61) | 1.77 (1.22) | WNL (range= 0.4-4.0 mIU/mL) | .961 |
| FT<sub>3</sub> (pg/dL) | 2.90 (0.26) | 2.58 (0.28) | WNL (range = 2.3-4.2 pg/dL) | **.021** |
| FT<sub>4</sub> (ng/dL) | 1.08 (0.17) | 1.17 (0.18) | WNL (range = 0.6-1.6 ng/dL) | **< .001** |
| rT<sub>3</sub> (ng/dL) | 18.8 (5.50) | 20.4 (5.32) | WNL (range = 9.2-24.1 ng/dL) | .178 |
| FT<sub>3</sub>:rT<sub>3</sub> | 0.17 (0.05) | 0.14 (0.05) | **OUTSIDE RANGE** (range= FT<sub>3</sub>:rT<sub>3</sub> ≥ 0.20) | **.004** |
| Hypothyroid symptoms (count) | 4.62 (3.67) | 5.71 (4.15) | No reference range | **.020** |

<sup>#</sup>Early post-surgery refers to 2 weeks to 3 months post-surgery.
Note: TSH = thyroid stimulating hormone; FT<sub>3</sub> = free triiodothyronine; FT<sub>4</sub> = free l-thyroxine; rT<sub>3</sub> = reverse 3,3',5'-triiodothyronine; FT<sub>3</sub>:rT<sub>3</sub> = ratio of FT<sub>3</sub> to rT<sub>3</sub>; **Bold** = significant $p < .05$

**Table 4.** Linear regression testing predictive associations between pre- to early post-surgery changes in the FT_3_:rT_3_ ratio and other exploratory variables associated with pre- to 1- or 2-years post-surgery changes in Body Max Index (BMI).

| Model | R<sup>2</sup> | Adj R<sup>2</sup> | Partial R | β | b (SE) | p |
| --- | --- | --- | --- | --- | --- | --- |
| **Change in BMI pre- to 1-year post-surgery (n=16)** |  |  |  |  |  |  |
| <u>**FT<sub>3</sub>:rT<sub>3</sub>**</u><br>↓ FT<sub>3</sub>:rT<sub>3</sub> | 0.58 | 0.52 | 0.45 | 0.47 | 15.67 (6.22) | **.026** |
| <u>**TSH**</u><br>↓ TSH | 0.48 | 0.39 | -0.40 | -0.32 | -5.71 (3.64) | .141 |
| <u>**FT<sub>3</sub>**</u><br>↓ FT<sub>3</sub> | 0.53 | 0.45 | 0.41 | 0.31 | 4.65 (2.96) | .143 |
| <u>**FT<sub>4</sub>**</u><br>↑ FT<sub>4</sub> | 0.42 | 0.33 | -0.25 | -0.21 | -4.93 (5.29) | .368 |
| <u>**rT<sub>3</sub>**</u><br>↑ rT<sub>3</sub> | 0.45 | 0.36 | -0.34 | -0.28 | -8.66 (6.72) | .220 |
| <u>**Hypothyroid symptoms (n = 12)**</u><br>↑Hypothyroid symptoms | 0.52 | 0.41 | 0.36 | -0.36 | -6.55 (5.74) | .284 |
| **Change in BMI pre- to 2 years post-surgery (n=13)** |  |  |  |  |  |  |
| <u>**FT<sub>3</sub>:rT<sub>3</sub>**</u><br>↓ FT<sub>3</sub>:rT<sub>3</sub> | 0.62 | 0.54 | 0.65 | 0.53 | 18.87 (7.04) | **.022** |
| <u>**TSH**</u><br>↓ TSH | 0.44 | 0.32 | -0.39 | -0.33 | -6.45 (4.86) | .214 |
| <u>**FT<sub>3</sub>**</u><br>↓ FT<sub>3</sub> | 0.43 | 0.31 | 0.23 | 0.18 | 3.76 (5.22) | .489 |
| <u>**FT<sub>4</sub>**</u><br>↑ FT<sub>4</sub> | 0.40 | 0.29 | -0.32 | -0.28 | -7.59 (7.20) | .317 |
| <u>**rT<sub>3</sub>**</u><br>↑ rT<sub>3</sub> | 0.52 | 0.42 | -0.52 | -0.45 | -15.45 (7.88) | .080 |
| <u>**Hypothyroid symptoms (n = 10)**</u><br>↑Hypothyroid symptoms | 0.35 | 0.17 | -0.39 | -0.36 | -7.14 (6.32) | .295 |

Note: All linear regressions included a surgery moderator variable as a covariate to control for effects of surgery type (i.e., RYGB or SG) on weight loss.
FT<sub>3</sub>:rT<sub>3</sub> refers to the ratio of free T<sub>3</sub> to reverse T<sub>3</sub>; TSH = thyroid stimulating hormone/thyrotropin; Hypothyroid Symptoms = total number of hypothyroid symptoms (out of 17 possible); FT<sub>3</sub> = free T<sub>3</sub>.
The dependent variable (DV) for the first set of models was the change in body mass index (BMI) from pre- to 1-year post-surgery. The DV for the second set of models was the change in BMI from pre- to 2 years post-surgery. The sample size at 2-year post-surgery is smaller than the 1-year sample due to patient attrition from follow-up between 1-year post-surgery and 2 years post-surgery (i.e., $n_{1\text{year}} = 16$ ; $n_{2\text{year}} = 13$ ). The availability of data at these time points for the hypothyroid symptom counts was less, with $n_{1\text{year}} = 12$ and $n_{2\text{year}} = 10$ . This is likely due to the fact that symptom count data required completing an online measure, a step beyond the request for the lab draws.

Notation is given as to whether the values for these markers over this period remained within the reference range or changed significantly.

Though there was a decline in the TSH from pre- to early post-surgery, the TSH remained in the normal range and the reduction was not significant. The FT_3_ also remained in the normal range, though its decline was significant. The FT_4_, which also remained in the reference range, increased significantly. The FT_3_:rT_3_ ratio, already below the reference range at baseline (as noted in the Discussion), showed a further significant reduction over this period. The value of the rT_3_ increased, though this increase did not move it outside the reference range and was not significant. The mean counts of self-reported hypothyroid symptoms increased from baseline to the early post-surgery period, with no reference range available.

#### Putative predictors of BMI from pre-surgery to 1-2 years post-surgery

The major outcome findings for the study are shown in Table 4. Only one thyroid measure significantly predicted weight loss outcome by linear regression at either 1- or 2-years post-surgery. Specifically, after controlling for the effects of surgery type (i.e., RYGB or SG) on weight loss, only decreases in the FT_3_:rT_3_ ratio from pre- to early post-surgery were significantly associated with a reduction in BMI (i.e., less weight loss) in the period between pre-surgery and both 1 year (*b* = 15.67, *SE* = 6.22, *p* = .03) and 2 years post-surgery (*b* = 18.87, *SE* = 7.04, *p* = .02). No significant effects were found for changes in other thyroid hormones (i.e., TSH, FT_3_, rT_3_, FT_4_) nor for total self-reported hypothyroid symptoms on weight loss outcomes at either time point (all *p*’s > .05).

## Discussion

This small exploratory study examined whether early indicators of the development of euthyroid sick syndrome (ESS) would predict later changes in weight loss among patients undergoing intentional weight loss via bariatric surgery. Our hypothesis received preliminary support in that early reductions in the FT_3_:rT_3_ ratio after bariatric surgery predicted significantly less weight loss at both 1- and 2-years post-surgery. No other marker of thyroid function (e.g., TSH levels, FT_3_, rT_3_, FT_4_, or hypothyroid symptoms) significantly predicted weight loss over that same period.

ESS refers to changes in thyroid hormones that typically arise following acute or severe illness that are not due to an intrinsic primary deficit in thyroid functioning [25, 26, 40, 41]. ESS patients are regarded as clinically euthyroid because of their normal TSH and T_4_ levels [25, 26, 40, 41]. To prevent excessive weight loss due to energy catabolism during the ill state, FT_3_ production is reduced and rT_3_ degradation is inhibited, thus lowering metabolism [40, 41]. This response is believed to be adaptive in ESS patients who are recovering from an illness, infection, trauma, or other stressors such as famine [25-33,36, 40-41]. As noted, reductions in the FT_3_:rT_3_ predict survival in patients with ESS including those undergoing the stress of surgery, severe illness, and infection [ 27-31]. However, for patients intentionally undergoing rapid weight loss in the context of bariatric surgery, suppressed metabolism can be considered maladaptive and counter to weight loss goals [42].

An important potential contribution of this study is that it addresses a common clinical dilemma of how to approach patients who report strict adherence to their post-bariatric hypocaloric food plan yet present with lower-than-expected weight losses (e.g., weight loss plateaus) [42]. In such situations, most clinicians might order a TSH level, particularly when the patient’s lack of expected weight loss is accompanied by symptoms suggestive of hypothyroidism (e.g., fatigue, cold intolerance, hair loss). Yet, if the TSH results are returned as within the normal or low-normal range indicating normal thyroid functioning (such as would be likely for patients in the current study), clinicians may be inclined to doubt the veracity of their patient’s self-report.

This small hypothesis-generating study offers a potential explanation to help understand why patients after bariatric surgery who are euthyroid by TSH might yet present with symptoms consistent with suppressed thyroid functioning (e.g., lack of weight loss despite hypocaloric intake accompanied by cold intolerance, fatigue). In such cases, the TSH may not be the most sensitive indicator of thyroid functioning post-surgery. Instead, the FT_3_:rT_3_ ratio post-surgery may be more useful for identifying the metabolic suppression characteristic of ESS.

While early changes in the ratio of FT_3_ to rT_3_ significantly predicted weight loss outcomes at 1- and 2-years post-years post-surgery, it is puzzling that only the numerator (FT_3_) showed a significant decrease over this early period. While rT_3_ levels in the denominator increased across this time frame, this rise did not reach significance. It is possible that the significant reduction in the ratio might be attributed to the FT_3_ alone or be the result of multiple comparisons within a small sample size. However, it is important to review the temporal nature of changes in rT_3_ levels. Prior studies show that following weight loss through diet or bariatric surgery, rT_3_ levels increase significantly by 2-3 weeks [8, 43] and regress towards baseline levels by 3 months post-surgery [8]. To reduce participant burden, the current study accepted rT_3_ labs collected at any time over the entire 2 week to 3-month post-surgery time period. Hence, it is possible that this wide range of post-surgical measurement time-points obscured the early significant increase in rT_3_. Additionally, because FT_3_ and rT_3_ exhibit opposing effects analogous to the gas (FT_3_) and brake (rT_3_) pedals of a car, the ratio of these two markers may be more sensitive in capturing these dynamic changes in thyroid functioning compared to either hormone alone, especially since only the ratio, and not the FT_3_ or rT_3_ levels were significant predictors.

Additional limitations are also important to note. First, this study included a small initial sample size (n = 23, of which n=20 had early post-surgical data available), with further losses at both 1- and 2-year follow-up. Hence, the analyses were subject to bias and underpowered to detect important differences. As indicated, the focus of this study was on our hypothesis that early development of ESS due to rapid weight loss in the early post-surgery period would predict reduced weight loss given the very low-calorie diets patients are asked to follow. These preliminary data should be considered exploratory in nature and useful only in that they offer intriguing initial support for our hypothesis. The small sample size would not be published as a standalone report but are intended as a small “proof of concept” accompanying our hypothesis. The suggestions from this study can help in designing larger investigations. Such future examinations are important given the poorly understood problem of suboptimal weight loss after bariatric surgery its considerable associated physical, emotional, and financial costs.

Unfortunately, the high loss to follow-up rate among post-bariatric patients is a notorious impediment to understanding the drivers of suboptimal outcomes. The attrition rates in this study, especially at the 2 year follow-up, were similar to those found in other bariatric follow-up studies [44-46]. Second, though our population was technically euthyroid at baseline based on TSH and FT_4_ values, and pre-surgical blood draws were purposefully obtained before patients began the required two-week liquid protein diet to reduce peri-operative complications, many patients likely already were following highly restrictive hypocaloric diets and already had developed ESS at “baseline.” Indeed, weight loss prior to surgery is often an insurance company mandate. The fact that further decreases in this ratio by early post-surgery predicted future weight loss, despite already abnormally low ratios at baseline, suggests the sensitivity of changes in this ratio as a prognostic indicator. Third, thyroid hormone labs were not collected at the 1- and 2-year follow-up time points. These data were not collected due this small hypothesis-driven study’s limited funds and would clearly be recommended in future studies. Of note, the early reductions in resting metabolic rate among the Biggest Loser participants (e.g., 610 ± 483 kcal/day below baseline at the end of the competition) persisted even when measured 6-years later (e.g., 704 ± 427 kcal/day below baseline) [47]. Unfortunately, the Biggest Loser study (just as previously mentioned POUNDS LOST study [18]) failed to include the rT_3_ in its thyroid test battery. Last, three of the 23 participants had diabetes (all type 2) at baseline. Each had documented resolution of diabetes symptoms within 2 weeks – 3 months post-surgery and only two of the three were included in the final analyses due to incomplete data. However, future studies should carefully consider the inclusion of participants with diabetes because of research showing a relationship between both diabetes and systemic inflammation with the development of ESS [48, 49].

To address these limitations and more definitively explore this study’s hypothesis of a predictive relationship between early changes in the ratio of T_3_:rT_3_ and later weight loss, we recommend future investigators: 1) recruit larger sample sizes; 2) collect thyroid hormones earlier in the pre-surgery process, such as at the first consultation (i.e., prior to the initiation of pre-surgery weight loss); 3) collect thyroid samples at post-surgery at both 2-weeks <u>and</u> 3 months; 4) exclude (or include as a separate arm) patients with diabetes; 5) check for thyroid autoimmunity and control for hypertension, dyslipidemia, medications and other factors that may influence thyroid metabolism ; and 6) measure weight, thyroid measures (including additional ratios, e.g., FT_4_/FT_3_) and other labs (e.g., c-reactive protein, fasting glucose, HgA1_c_) at frequent intervals throughout the follow-up period. Such measurements will enable a more robust testing of the hypothesis as well as allow clearer distinction between weight loss and weight regain trajectories.

The above recommendations are meant to help future researchers design needed studies to further investigate the hypothesis that the FT_3_:rT_3_ ratio is a useful marker of weight loss outcomes in bariatric patients, as well as in other individuals who experience weight loss plateaus and symptoms of hypothyroidism (e.g., fatigue, hair loss, cold intolerance) despite strict adherence to hypocaloric diets.

Clinical implications of this hypothesis generating study, if replicated, include the possibility that the FT_3_:rT_3_ ratio can potentially serve as an early predictor of reduced weight loss outcomes in early post-bariatric patients experiencing hypothyroid symptoms in the absence of changes in TSH. Indeed, there may be a window within the early post-surgery period during which reductions in FT_3_:rT_3_ ratio may not only identify patients at-risk for longer term poorer weight loss outcomes, but offer an opportunity for clinical intervention (e.g., potential repletion of T_3_) [50].

## Conclusions

Given the high percent (up to 30%) of patients with suboptimal weight loss outcomes after bariatric surgery, identification of early prognostic indicators of later weight loss is important. The paper is the first of which we are aware to investigate whether early development of ESS symptoms would predict later post-bariatric weight loss. The finding that early reductions in the FT3:rT3 ratio predicted reduced weight loss outcomes at 1 and 2 years after bariatric surgery provides valuable but very preliminary data to be used for further hypothesis generation and future study design, while being interpreted with caution due to the limitations noted. Investigations to further advance our understanding of predictors leading to optimal or suboptimal bariatric surgery weight loss outcomes are needed. If the FT3:rT3 ratio is shown to have greater sensitivity than the TSH in identifying suppressed thyroid metabolism early post-bariatric surgery, there may open a useful window within the early post-surgery period to maximize outcomes through early identification and intervention [50].

## Data Availability

All data produced in the present study are available upon reasonable request to the authors.

## Acknowledgements

The authors would like to thank Courtney Crisp for her contribution in recruiting participants for this study.

